# Genome-wide association study of John Henryism in the CARDIA Cohort suggests a molecular mechanism behind lower nicotine addiction rates

**DOI:** 10.1101/2023.05.18.23289991

**Authors:** Richard R Chapleau

## Abstract

John Henryism (JH) is a stress response mechanism that enables individuals to cope with chronic psychological stressors. Originally identified in African American males, JH has since been associated with numerous negative health outcomes such as cardiovascular disease. Despite its relationship to diseases with known genetic risk factors, little work has been reported concerning the genetics of JH. In the current study, genome-wide microarray and the JH Active Coping scale data from the CARDIA Cohort study were used to identify genetic factors associated with JH levels. Principal component analysis accounted for population stratification and evaluated six inheritance models in plink software. We also performed network analyses on the resulting significant associations (P < 5×10^−8^) to identify molecular pathways to health outcomes. Our GWAS results revealed 25 significant genetic associations and two suggestive associations. One of the variants identified with a suggestive association (P < 5×10^−6^) to JH (rs11634680) reproduces the same suggestive association from a prior study of the same dataset with the similar odds ratios and P-values between the studies. In our pathway analysis, we found a variant associated with JH (rs12448959) decreases the amount of GABA transaminase and thereby reduces the nicotine reward through the GABAergic signaling pathway. Our work provides a molecular explanation to the observational data that individuals with high levels of JH active coping skills are less prone to using smoking as stress relief.

## INTRODUCTION

John Henryism (JH) is a psychological construct that denotes a strong behavioral tendency to cope actively with chronic stressors such as racial discrimination and economic challenges by exerting high levels of effort, which entails accumulating physiological costs [1]. The effects of JH manifest in both clinical and non-clinical settings, affect emotional responses and may influence the functioning of individuals, especially African Americans, in their daily lives [2]. Over time, the accumulated stress can precipitate the early onset of cardiovascular disease [3]. The negative relationship between JH and cardiovascular health is also reported with respect to income levels, where hypertension is more prevalent in low-income individuals scoring high in JH [4]. This suggests that the addition of a financial stressor induces JH active coping, resulting in increased physiological damage. The use of increased effort as a coping strategy, especially in the face of discrimination, could potentially backfire when considering a holistic health viewpoint.

While evidence suggests that high levels of JH active coping negatively impact physical health, the relationship between JH and mental health appears to be largely protective. For instance, in Black women it appears that there is a negative correlation between JH and symptoms of depression [5]. Additionally, in Black Americans of low socioeconomic status, high levels of JH have been linked to improved mental health [6]. Higher levels of JH active coping were also associated with lower odds of needing mental health services or reporting opioid abuse [7]. JH also moderates the increasing effect of racial discrimination on depressive symptoms [8]. Collectively, the research evidence appears to suggest that JH active coping is beneficial for the mental health of individuals responding to stresses applied through social and cultural infrastructure.

We hypothesized that there are genetic associations with JH active coping as this social construct is passed through generations [9], and the heritability of JH has been reported at more than 30% in two twin studies [10, 11]. Here, we report the results of our study specifically investigating genetic associations with JH. Following on a previous genome-wide association study (GWAS) which used a test and validation approach [12], we attempted to evaluate the effect of population stratification on genetic associations. In contrast to the prior approach, here we included all samples as a single cohort specifically testing for JH.

## MATERIALS AND METHODS

Our study was approved by the WIRB-Copernicus Group Institutional Review Board (Study number 1332892) and we followed the Helsinki declaration. We used data from the National Institutes of Health’s Database of Genotypes and Phenotypes (dbGAP). The data came from the Coronary Artery Risk Development in Young Adults (CARDIA) Study (dbGAP accession phs000285) [13]. We followed the STREGA guidelines for genetic association studies from the EQUATOR network [14].

### Phenotype and Genotype Data pre-processing

JH was measured by the 12-item John Henryism Active Coping Scale (dbGAP accession number phv001133534.v2.p2.c1), and responses were reverse-coded. The JH score was calculated as a mean of the responses. We also defined a binary “high JH” variable, categorizing individuals with scores above the median as “high” [2,3]. We replaced missing data with the average. We obtained genotype data from two CARDIA sub-studies, which used the Affymetrix Human SNP-6 microarray chips. We merged and filtered the data for autosomal genotypes. We only kept genotypes with less than 10% missing data (sample or locus), a minor allele frequency above 1%, and a Hardy-Weinberg equilibrium above .0001. The final sample size was 2,317. We used plink1.9 [15] for genetic pre-processing and R version 4.2.0 [16] for phenotype processing.

### Genome-wide association

We performed our GWAS using plink 1.9 [15]. We used the total scores (not the individual questionnaire responses) and the binary classification as outcomes. We ran four tests (using the --linear recessive/genotypic/hethom/dominant flags for plink) for each outcome, which tests six distinct inheritance models (additive, dominant, recessive, genotypic, dominance deviation, heterozygotic, and homozygotic). We used linear or logistic models depending on the outcome, selected automatically by plink. To avoid population stratification, we also performed principal component analysis (PCA) in plink. We repeated the GWAS approach using the six inheritance models, using the first four principal components (PC) as covariates.

### Gene annotations and network analysis

We used SNPtracker [17] and FAVOR v2.0 [18] to find gene and variant names for the significant associations. The dbSNP (Database of Single Nucleotide Polymorphisms) Batch Query used GRCh38, dbSNP build 155. The FAVOR annotations used GRCh38 and the Trans-Omics for Precision Medicine Freeze 8 BRAVO variant set [19]. We used STRING (www.string-db.org) [20] to predict interaction networks. We used a minimum association score of 0.150, no more than 5 first shell interactions for less than 20 proteins, and no second shell interactions for any number of proteins.

## RESULTS

The dataset properties have been reported in detail elsewhere [12]. Briefly, the JH score had a median of 49 and a high JH category with 1497 observations. The sex ratio was slightly imbalanced (42% male). The variables deviated from normality (Shapiro-Wilk’s W = 0.980 for JH scores and 0.635 for high JH), exhibited negative skewness (−0.511), and had low kurtosis (k = 3.11).

### Unadjusted GWAS Results

Without adjusting for population stratification, we detected more than 40 GWAS-significant (P < 5×10^−8^) associations in an allelic/additive inheritance model (Figure 1), consistent with prior reports [12]. Dominance deviation was the only model that did not show a significant association with JH score, and all models except homozygous and recessive showed significant associations with high JH. The large number of significant associations (up to 704 in the genotypic model) may indicate a population stratification confounder.

**Figure 1:**
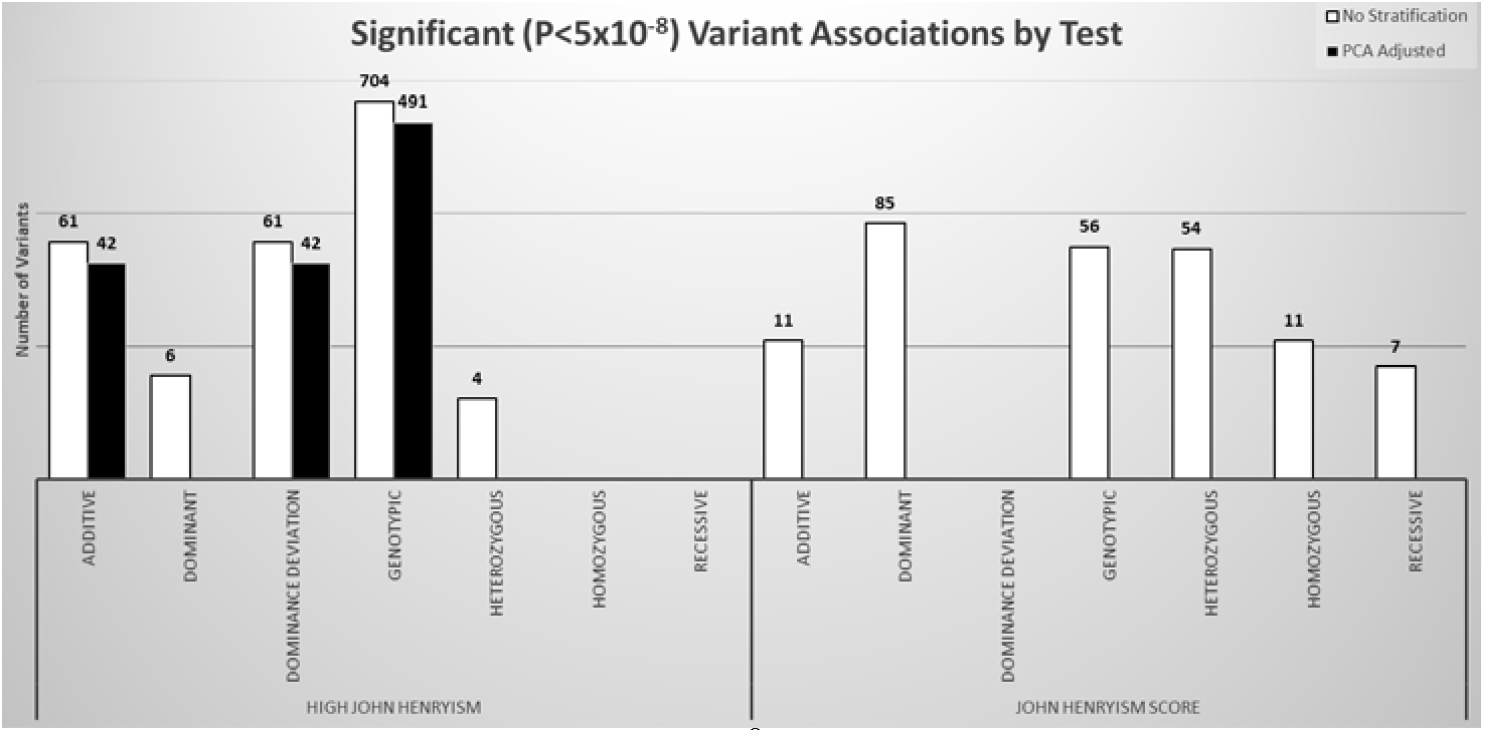
Count of GWAS significant (P<5×10^−8^) variants associated with John Henryism Active Coping score or high John Henryism. Significant associations were identified in most inheritance models when not accounting for population stratification (white columns). However, when stratification is accounted for (black columns), none of the quantitative trait analyses showed significant associations and the binary trait analyses identified fewer significant associations.

### PCA-Adjusted GWAS Results

The PCA analysis revealed that the first PC explained 74.9% of the variance in the dataset (Supplementary Figure S1). Moreover, the first four PCs accounted for 81.3% of the variance. Additionally, the variance explained plot showed that a linear trend fitted to the full dataset had a lower R^2^ value (0.989) than a linear trend fitted to the first four PCs and the remaining 16 PCs (0.999 and 0.999, respectively). By adjusting for the first four PCs as covariates, we detected 42 GWAS-significant (P < 5×10^−8^) associations for high JH in the PCA-adjusted GWAS using an additive model (Figure 2). The same 42 associations were detected with the dominance deviation inheritance model.

**Figure 2:**
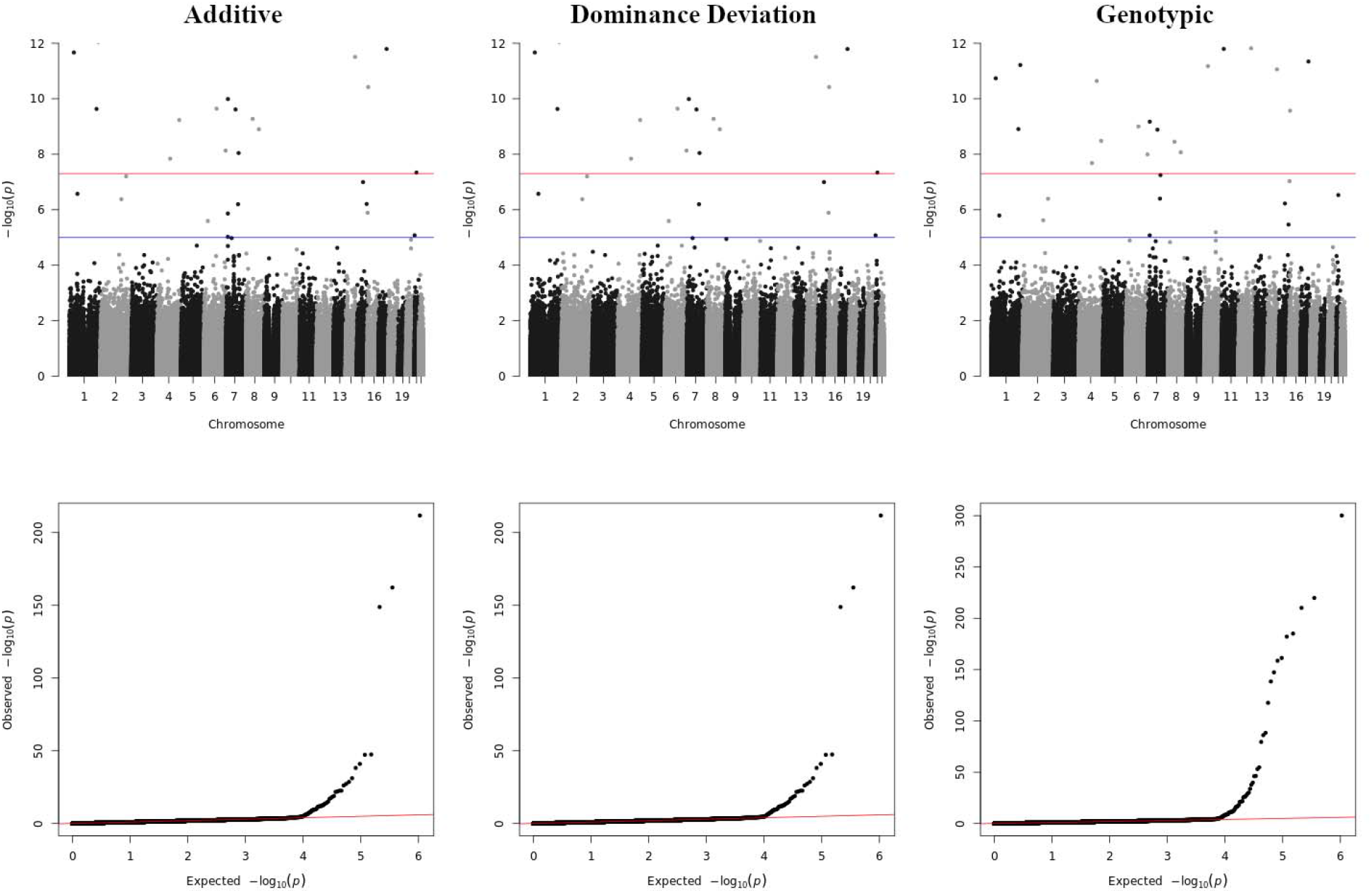
Manhattan and qq plots of association with high John Henryism for the three inheritance models reporting genome-wide significant associations (P<5×10^−8^).

Of the 42 significant associations, 25 variants were returned from the plink analysis with an odds ratio (OR) of “Inf”, suggesting an arithmetic overload possibly caused by low allele frequencies. An additional 17 were calculated with an OR of 0 and a large standard error and are not reported herein. Of the 51 variants that met the GWAS “suggestive” threshold (P<5×10^−6^), only 2 had finite ORs calculated by plink and are reported in Table 1 alongside the 25 “significant” associations with infinite ORs. Interestingly, the genotypic model yielded more than 10-fold increase in the number of significantly associated variants (491 associations), but ORs are not calculated for the genotypic model.

**Table 1:**
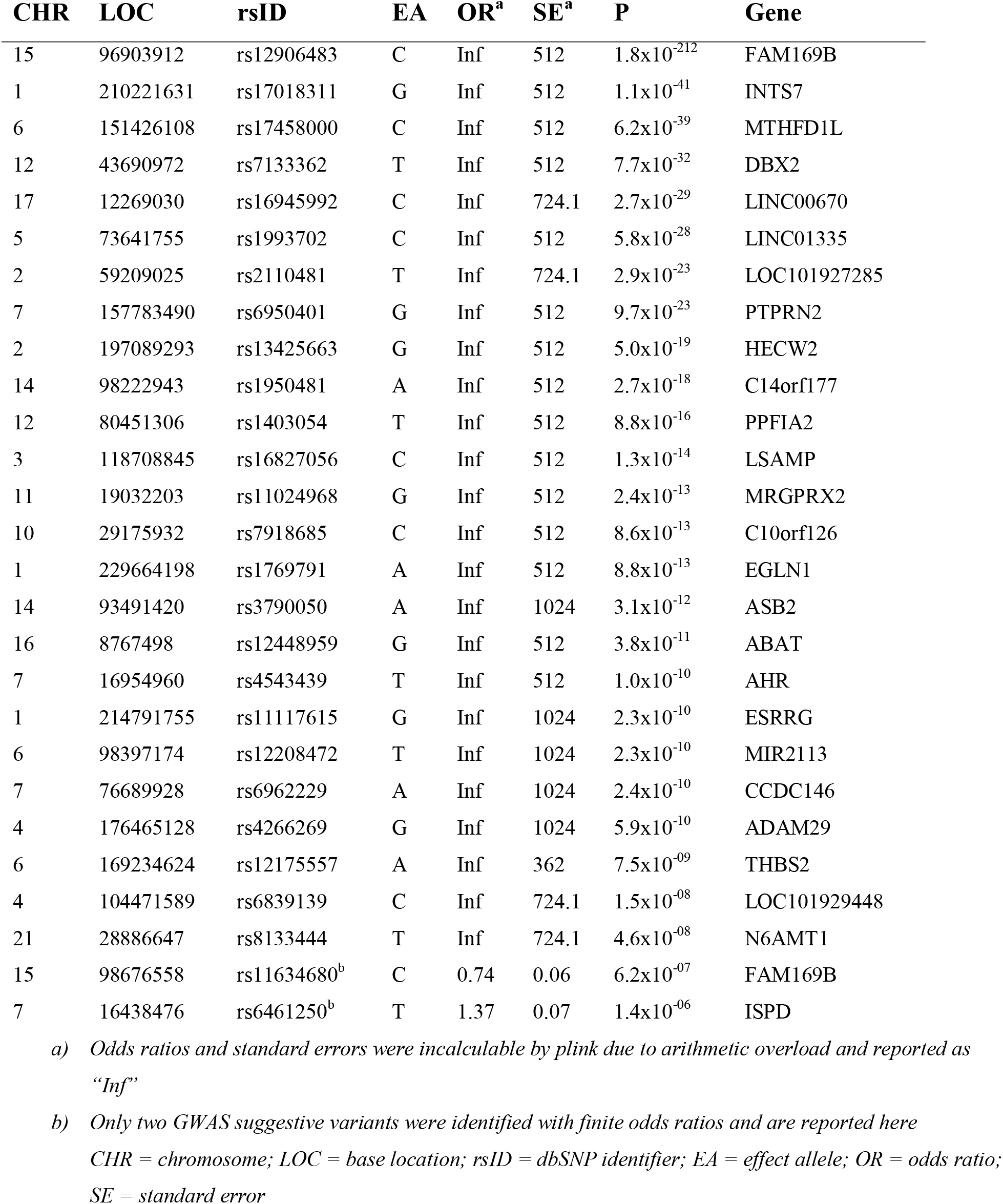
Summary statistics for PCA-adjusted variant associations.

### Network Analysis on PCA-Adjusted Significant SNPs

We identified 17 associated genes from the PCA-adjusted additive GWAS results and 208 associated genes from the genotypic GWAS and used those genes lists as the input for network analysis using the STRINGdb web resource. As the additive model had fewer than 20 proteins in the query list, we included an additional 5 first order interacting proteins in the search criteria. Our additive GWAS network contained 22 nodes and 41 edges (Figure 3A) with a protein-protein interaction enrichment P-value of 0.026, indicating a significantly increased number of interactions as compared to the number of interactions expected from a random set of 22 genes (expected value = 29). Similarly, for the 208 genes associated in the genotypic model results, we found 1,324 edge interactions compared with an expected 1,085 interactions (P = 1.28×10^−12^). Using the STRINGdb resource to search pathway and publication databases, the networks we found in the additive GWAS results accounted for 72 enriched terms in 8 distinct databases (Supplemental Table S1), while the genotypic networks accounted for 126 enriched terms in 6 databases (Supplemental Table S2). None of the terms were matched between the two lists. The enriched terms from the additive results primarily represent underlying biological mechanisms and pathways, whereas the terms enriched in the genotypic results are primarily derived from self-reported health outcome data and terms reported in GWAS databases.

**Figure 3:**
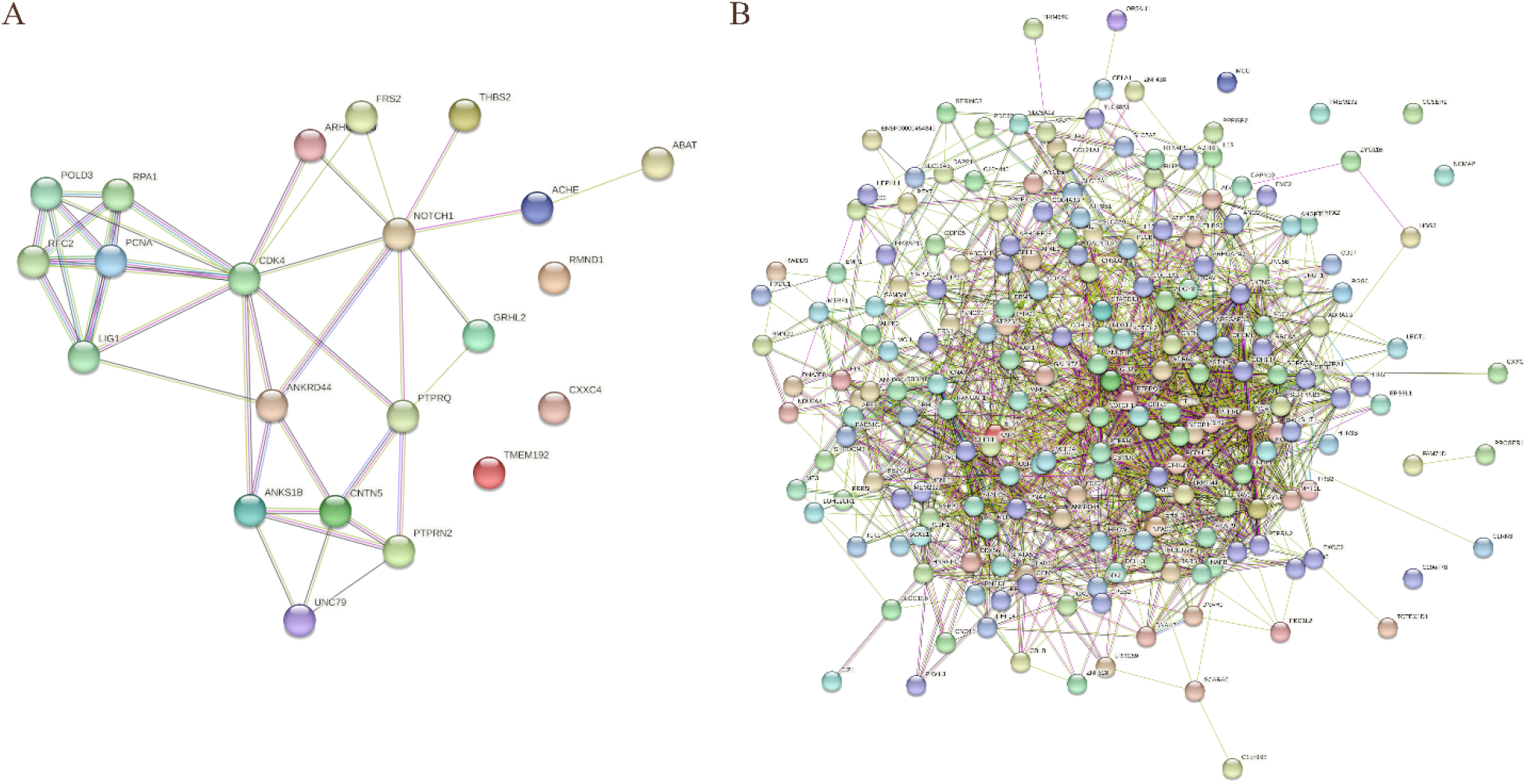
Network diagrams for genes significantly associated with John Henryism in the additive (A) or genotypic model (B). The additive model produced fewer nodes than 20 nodes and therefore include 5 first-order network connections.

## DISCUSSION

The genetic analyses indicated a high level of population stratification in the CARDIA cohort. With the two-stage approach taken in our prior study, the sample size of our discovery stage was 1,852 participants after data quality control. In the current study, where we used all data jointly, we had 2,317 participants which is a net increase of 25.1%. In the prior study, we detected 3 candidate genetic variants associated with JH. Without adjusting for population stratification in our current study, we detected 61 variants in the additive model (Figure 1), none of which were replicated in our current study. Our PCA adjustment for population stratification reduced the number of associated variants to 25 significant associations with JH and two additional suggestive associations (Table 1). One of the variants identified here as suggestive (rs11634680) was also identified as suggestive in the first study that evaluated the associations between JH and cynicism [12]. In this study, we identified an OR of 0.74 with an error of 0.06 (95% confidence interval: 0.61 to 0.86) and a P-value of association of 6.2×10^−7^. In the prior study, the OR was 0.71 with a P-value of 3.3×10^−7^. As both the P-value and the OR are similar, we conclude that we were able to successfully replicate this association from the same data set.

Misspecification of genetic models in GWAS can increase the risk of false negatives, but multiple model testing can also increase the risk of false positives [20-22]. Given the small effect sizes and common variants expected for these traits, we chose to test multiple inheritance models. All variants detected in the additive models were also detected in the dominance deviation models, suggesting no dominance effect of one allele over the other, so we only report the differences between the additive and genotypic models. The genotypic model does not assume any inheritance pattern, and therefore was likely to have a large number of variants, many of which may not be replicated in more specific models or additional testing.

Higher levels of JH have been associated with lower levels of smoking [21], though the mechanism behind this association is unknown. Our results provide insight into one possible biological pathway. We found an association between a T-to-G transversion in an H3K4me1 enhancer located upstream of the *ABAT* gene (rs12448959), where the G variant resulted in a decreased odds of experiencing high JH. These monomethylated enhancers can act as alternate promoters in some tissues [22], but primarily function by delivering accessory transcription factors (TF) to the promoter [23]. Transversions (Tv) are typically found to interrupt TF binding [24], and in our case the mutation found in individuals with high JH results in increased GC content. Therefore, one potential pathway to increased smoking rates and nicotine addiction in individuals with low JH is that the *ABAT* Tv we found results in increased levels of ABAT protein [25-27], which catabolizes the neurotransmitter GABA [28], which in turn means a decrease in available GABA for binding to the GABA_B_ receptor resulting in an increase in the nicotine reward [29]. Taken together, individuals with high JH appear to experience less reward from nicotine use and would subsequently be not as susceptible to addiction.

## CONCLUSION

We presented our findings from a GWAS for JH. By adjusting for different inheritance models and population stratification, we detected 25 significant associations albeit without calculable ORs and we detected two suggestive associations with finite ORs, one of which was also present in a prior study on this data set. We also conducted an unadjusted GWAS to replicate previously published results using a two-stage discovery and validation approach. The results of the current study support the conclusions of the first study that there is a genetic component to the development of JH. We also developed potential molecular mechanisms supporting observational evidence connecting the JH active coping strategy with reduced reliance upon nicotine for tolerating psychological stress.

## Data Availability

Genotype and phenotype data that support this study can be obtained from dbGAP using accession numbers phs000285.v3.p2, pht001579.v2.p2, pht001634v2.p2, pht001786.v2.p2, phg000092.v2, and phg000098.v2. GWAS summary statistics based created from GWAS on the training dataset are unavailable for public posting, per the CARDIA data use agreement. The summary statistics are available from the author upon reasonable request.

## ACKNOWLEDGMENTS

We are grateful to Mr. Billy Thompson, Ms. CharLee Martin, Mr. Eric Rigby, and Mr. Warren Fridy for their help in arranging data access and security. We also thank Dr. Tyler Mulhearn and Ms. Tanya Goodman for their critical comments on the manuscript. We also appreciate the participants and the original contributors of the CARDIA study (phs000285).

## FOOTNOTES

### Institutional review board statement

This study was reviewed and approved by WIRB-Copernicus Group Institutional Review Board (Study number 1332892).

### Data sharing statement

The authors have read the STREGA guidelines for reporting genetic association studies and prepared the manuscript accordingly.

